# Evaluating the Gut Microbiome, Dietary Patterns, & Cognition: A Sub-study Protocol from the Brain Health & the Gut Microbiome Study in Cognitive Decline (bMicrobiome Study)

**DOI:** 10.64898/2026.02.02.26345243

**Authors:** Karol Suchowiecki, Patrick G. Corr, Aidan Schurr, Aryan Asemani, Leigh A. Frame

**Affiliations:** Department of Medicine, University of Connecticut Health Center; The George Washington University School of Medicine and Health Sciences; The George Washington University School of Engineering and Applied Sciences; The George Washington University Milken Institute School of Public Health

**Author notes:** **Corresponding Author:** Karol Suchowiecki, 263 Farmington Ave Farmingtonm CT 06030.

**Keywords:** gut microbiome, dietary patterns, cognitive decline, Alzheimer’s disease, behavior change, microbiome composition, dietary assessment

## Abstract

**Objectives:** To investigate how nutrition readiness to change influences implementation of dietary behavior changes and to compare the gut microbiomes and document gut microbiome composition changes over time in individuals with early Alzheimer’s disease dementia (eAD), mild cognitive impairment (MCI), and healthy controls (HC) Overall, this study aims to add to the emerging field of how the gut microbiome influences the nervous system.

**Methods:** This is a sub-study of a multi-prong proof-of-concept, observational study mapping the gut microbiome: 15 HC, 15 MCI, 15 eAD (n=45). At 0-, 3-, and 6-months, participants are provided lifestyle recommendations tailored to their gut microbiome. Participants may choose to implement this or not and are observed throughout (observational intervention study). In this sub-study, a survey is developed and implemented in conjunction with dietary assessment (DietID) to evaluate the role of Readiness to Change in implementation of dietary recommendations.

**Results:** This is the sub-study protocol from an ongoing parent study.

**Discussion:** This protocol presents a novel intervention to assess the gut microbiome, individual dietary patterns, and readiness to make lifestyle change related to diet.

**Trial Registration:** NCT06039267

## Background

### The Gut Microbiome and Cognitive Health

The gut microbiome is composed of trillions of microorganisms (microbiota) that play a vital role in human health and pathogenesis of disease, particularly as related to cognitive function and Alzheimer’s disease and related disorders (ADRD) (Guinane & Cotter, 2013; Manderino et al., 2017). Research has established that there is a possible protective factor against cognitive decline in the gut microbiota (Harding & Bishop, 2022; Liang et al., 2022). While we cannot define what a “healthy” gut microbiome is, low microbiota diversity is a common trait of an “unhealthy” gut microbiome also commonly referred to as “dysbiosis.” Animal studies and small clinical trials are establishing a possible relationship between symptomatic ADRD and gut dysbiosis (Quigley, 2017). A recent cohort study by Ferreiro et al. demonstrated that individuals with preclinical Alzheimer’s disease had distinct gut microbiota composition and that change in composition correlated with β-amyloid (Aβ) and tau pathological biomarkers (Ferreiro et al., 2023). Presence of oral bacteria have been found in the postmortem brains of persons with Alzheimer’s disease (Hashimoto, 2023). Dysbiosis is thought to cause intestinal permeability termed “leaky gut,” which leads to a local inflammatory response. This local release of cytokines and subsequent immune stimulation can, in turn, stimulate a systemic inflammatory response that alters the blood-brain barrier and leads to neuroinflammation, which can result in neuronal and axonal injury over time (Quigley, 2017). Communication between the gut and brain occurs via direct and indirect signaling. Direct communication occurs through the vagus nerve while indirect signaling is thought to occur through the enteric nervous system (ENS), autonomic nervous system (ANS), central nervous system (CNS), neuroendocrine system, tryptophan metabolism, and microbial-derived metabolites (Warren et al., 2024). Dietary patterns can influence crosstalk between the gut and brain through both direct and indirect signaling via mechanisms such as tryptophan metabolism and production of microbial metabolites (Hirschberg et al., 2019; Sinha et al., 2024).

### Dietary Patterns

Individual dietary patterns directly influence our gut microbiome composition as these microbes rely on extracting nutrients from our digested food to survive. Bacteria in the microbiome utilize microbiota accessible carbohydrates (MACs) such as resistant starch and fiber for fuel. In this process they produce secondary metabolites such as short-chain fatty acids (SCFAs), secondary bile acids (SBAs), tryptophan metabolites like serotonin, glucagon-like peptide-1 (GLP-1), peptide YY (PYY), and numerous others, which have an effect on the gut-brain axis (Warren et al., 2024). Diets rich in MACs correlate with the production of SCFAs in the gut (Ayakdaş & Ağagündüz, 2023). Likewise, production of SBAs by gut microbiota is affected by dietary fat and fiber intake. Dietary fiber binds to SBAs in the gut and prevents their reabsorption, thus increasing gut concentrations; dietary fat is utilized in the production of cholesterol from which bile acids are synthesized (Wolf et al., 2022). Changes in SBAs and SCFAs concentrations are associated with gut barrier function and, thus, may result in leaky gut and increased intestinal inflammation (Warren et al., 2024). SCFAs are known to cross the blood brain barrier (BBB) and, therefore, are thought to play a role in microbiota-gut-brain crosstalk (Silva et al., 2020). Concentrations of these metabolites produced by the gut microbiota likely play a role in numerous disease states including Alzheimer’s disease (Silva et al., 2020). Different dietary patterns lead to varying concentrations of microbiota-derived metabolites, which may significantly influence a wide range of disease states and contribute to dysbiosis or a balanced gut microbiome.

Diets such as the Mediterranean diet (MedDiet) have been shown to change microbiome composition over time (Ticinesi et al., 2024). Short-term (2-4 week) MedDiet intervention studies found an increase in diversity of bacterial species (gut microbiome diversity) from baseline (Barber et al., 2021; Godny et al., 2022; Rejeski et al., 2022). That said, any regular diet will impact the composition of the gut microbiome; in other words, eating has the greatest effect on the makeup of our gut microbiome (Zhang, 2022). Diets like the MedDiet are rich in vegetables, fruits, nuts, beans, olive oil, and fish are high in fiber, mono- and poly-unsaturated fatty acids (MUFA/PUFA), antioxidants, and polyphenols (Nagpal et al., 2019). The fermentation of these dietary components results in production of specific metabolites such as SCFAs (Khavandegar, 2024). A systematic review with five observational studies reported a significant increase in SCFAs following MedDiet adherence (Khavandegar, 2024). It is evident that dietary changes lead to changes in microbiota composition and production of microbiota-derived metabolites, which are associated with numerous disease states including brain-related disorders like Alzheimer’s disease.

### Readiness for Change

Changing dietary patterns requires behavioral change, a challenge in addressing individual health (Sutton, 2022). The Transtheoretical Model of Change (TTMC) is a well-established framework to describe and categorize the dynamics of individual health-related behavioral change (Prochaska & DiClemente, 1992; Prochaska & Volicer, 1997). This model emphasizes five phases: precontemplation, contemplation, preparation, action, and maintenance. TTMC was first used in research related to psychotherapy, addiction, smoking cessation, weight loss, and alcohol and substance abuse (Schaffer, 2013).

The first stage of change is precontemplation in which there is no intention to make a change in a particular behavior at least in the next six months (Shaffer, 2013). People in this stage of change either do not know that they have a problem that needs change, or they may have tried to change in the past but failed and now have given up. Usually, this group of people is in denial that they have a problem and are even willing to argue that their behavior is not an issue (Raihan et al., 2023). The second stage of change is contemplation in which people are willing to make a change in the next 6 months. This stage is characterized by awareness that a problematic behavior exists and the individual is seriously considering change; however, there is uncertainty as to whether the behavior is worth changing (Raihan et al., 2023). The third stage of change is preparation and is characterized by intention to make a change in the near future, often within a month (Prochaska & Volicer, 1997). In this stage, people have determined that the pros outweigh the cons of the change, and they are usually obtaining vital information to make the change such as self-help resources and professional advice. The fourth stage of change is action, which occurs when people make a specific and overt change in their life within the previous 6 months (Prochaska & Volicer, 1997). Typically, a change in this category needs to meet a certain standard to reduce disease risk. For example, in a person trying to quit smoking a change to smoking no cigarettes would be adequate whereas a change to smoke 1-2 fewer cigarettes would not amount to a substantial enough change. Lastly, maintenance is the final stage of change where people have made a change and stuck to it for at least 6 months. They are usually working to prevent relapse, but they do not have to use as much effort to maintain the change as people in the action stage (Raihan & Cogburn, 2023). Some researchers include “termination” as the last stage of change; however, it is very hard to achieve and often there is always minimal effort required to maintain change, placing most people in the maintenance phase forever. Although the process of change is often not linear, and people often jump in and out of stages, the TTMC provides a valuable framework for behavioral change both in the clinical and research settings.

Numerous validated surveys have been developed based on this model including a 12-question readiness to change survey and a 32-question University of Rhode Island Change Assessment (URICA) (Heather et al., 1993; Henderson et al., 2004; McConnaughy et al., 1983). Although many iterations of the surveys exist, most have focused on addition to smoking, drugs, alcohol, or readiness to start psychotherapy. There was no previously published readiness to change survey that sought to evaluate nutritional and dietary change based on the TTMC at the onset of this study.

### Study Objectives

This is a sub-study of a larger study seeking to map participant microbiomes to establish gut composition among a population of healthy controls, participants with mild cognitive impairment, and participants with early Alzheimer’s Disease dementia. This study specifically seeks to explore the relationship between the gut microbiome and dietary patterns, ultimately to identify educational protocols to encourage individual behavior change and improve cognitive health.

### Main Study Objective

1. To compare the gut microbiomes of patients with early Alzheimer’s disease dementia, mild cognitive impairment, and healthy controls using diversity as well as genus, species, and strain level differences in composition and function.
2. To document microbiome changes following lifestyle changes in subjects with early Alzheimer’s disease dementia, mild cognitive impairment, and healthy controls

### Sub-study Objective

To investigate how participants’ nutrition readiness to change influences their implementation of dietary behavior changes and the subsequent impact on study outcomes, particularly in relation to gut microbiome composition and cognitive health markers among individuals with early Alzheimer’s disease dementia, mild cognitive impairment, and healthy controls.

## Methods

### Study Participants

This sub-study recruited males and females stratified into three distinct groups as evaluated by a geriatrician with expertise in cognitive and gut health: healthy controls (n=15), those with mild cognitive impairment (n=15), and those with early Alzheimer’s disease dementia (n=15). Inclusion criteria included age 50-90, diagnosis of early Alzheimer’s disease dementia, diagnosis of mild cognitive impairment, and healthy controls. There were no specific exclusion criteria, aside from participants not meeting the inclusion criteria. Healthy controls were not screened for subjective cognitive decline. The inclusion and exclusion criteria for this study are broad and exploratory given that this is an early stage feasibility study. Details are reported under the ClinicalTrials.gov registration for the parent study (bMicrobiome Study, NCT06039267): https://clinicaltrials.gov/study/NCT06039267.

### Recruitment

Study participants were recruited from around the Washington, DC Metro Area, particularly from an integrative medicine clinic, memory clinic, and general internal medicine clinic which are all affiliated with a large academic medical center in DC. Potential study participants self-identified to be contacted by the study team. Each participant was provided an oral explanation of the study and written consent to review. Concerns and questions were addressed. The potential participants were made aware this study is entirely voluntary and that they could end their participation at any point. Participants who decided to enroll gave consent remotely, as per the decentralized design of the parent study. As study procedures required the completion of online surveys and remote assessments, access to a digital, internet capable device (e.g., a smartphone, tablet, or computer) was an inclusion criteria. Of note, for participants with early Alzheimer’s Disease dementia, caregivers are to be involved in the process of data collection, specifically to understand their role in influencing dietary patterns. These contextual factors will be considered in data analysis and may be presented as a possible limitation to the dataset.

### Study Design

This is an observational, sub-study of a proof-of-concept study mapping the gut microbiome of the study participants (previously described). This sub-study included three distinct data collection methods. For this sub-study, we gathered data on gut microbiome composition (EzBiome),the Boston Cognitive Assessment (BOCA), individual dietary patterns (DietID), and Readiness for Change stage (REDcap survey). These methods are described in detail below and a timeline of data collection is present in Table 1.

**Table 1:** Data Collection Process & Alignment.

| Activity | Baseline | Month 3 | Month 6 |
| --- | --- | --- | --- |
| Gut Microbiome Profiling | X | X | X |
| Dietary Assessment | X | X | X |
| Readiness for Change Survey | X | X | X |
| Boston Cognitive Assessment (BOCA) | X | X | X |

### Boston Cognitive Assessment (BOCA)

The BOCA is a self-administered computerized cognitive test developed for longitudinal cognitive monitoring. BOCA evaluated multiple domains of cognition including attention, memory, language use, executive function, visuospatial ability, and orientation (yshedskiy, 2022). In this study, BOCA is used to measure cognitive function across multiple time points (0-, 3-, and 6-months) and is typically completed in approximately 15-minutes on a table, computer, or smartphone. BOCA’s design minimizes learning effects by utilizing random, never-repeating tasks, making it suitable for repeated measurements over the study duration.

Participants access and complete the BOCA remotely through a secure, HIPAA-compliant platform. BOCA also provides a feasible, reliable cognitive function measure without necessitating in-person visits. Participants receive detailed instructions on accessing and completing the assessment online with study team assistance as needed. Members of the study team will be available to provide technical troubleshooting, step-by-step instructions, and real-time assistance in accessing the tool. Results are collected and stored securely, contributing to longitudinal data for each participant, which is essential for tracking potential cognitive changes over the study period. BOCA is complemented by additional self-report and caregiver assessments to enrich the data on participants’ cognitive and functional status.

### Gut Microbiome Profiling

Gut microbiome analysis in this study is conducted in an academia-industry-government partnership with EzBiome, the GW High-performance Integrated Virtual Environment (HIVE) Lab, and the National Institute of Standards and Technology (NIST), leveraging 16S rRNA sequencing and functional prediction to assess bacterial microbiome composition and function. For this analysis, a stool sample will be collected at home at baseline, 3 months, and 6 months with 2 consecutive samples to be integrated into a single EzBiome report at each time point. Instructions to the participants note that stool samples may be collected whenever they have a natural bowel movement (i.e., there is no prescribed time by which participants are required to collect their sample). The participant will receive their EzBiome reports and be advised to consider implementing the suggested lifestyle changes at each time point (baseline, 3 months, and 6 months). Any changes the participant makes (either recommended in the EzBiome report or otherwise) while participating in the study will be documented by the study team. At each time point, the stool samples will be analyzed, offering insights into microbial diversity and composition at the phylum, class, order, family, genus, and species levels as well as prediction of functional capabilities of the gut microbiota.

16S rRNA sequencing enables the identification of key bacterial taxa and patterns associated with cognitive health. For the researchers, the EzBiome report provides detailed data on the microbiome’s diversity index and specific microbial signatures linked to inflammation and gut health. For the participants, the EzBiome report provides tailored feedback based on their microbiome profiles, including personalized lifestyle recommendations intended to promote microbiome balance. These recommendations may include dietary adjustments and lifestyle modifications, with changes tracked to evaluate their impact on gut health and study outcomes over time.

All sample processing and analysis are conducted in a CLIA/CAP-certified laboratory with independent verification by NIST and the HIVE Lab, ensuring high standards for quality and data accuracy. Findings from each time point contribute to the study’s exploration of the microbiome-gut-brain axis, focusing on potential associations between microbiome composition, microbiome function, cognitive health, and participants’ readiness to make dietary changes.

### Dietary Assessment

DietID is a validated dietary assessment tool used to improve dietary management, streamline data collection, and support clinical optimization. A primary benefit of DietID is its modern, digital platform that has been shown to strongly correlate with common food frequency questionnaires (FFQs), 24-hour recalls, food records, and biomarkers (Bernstein et al., 2023; Dansiger et al., 2023; Katz et al., 2020; Radtke et al., 2023; Turner-McGrivey et al., 2021).

This makes the task of collecting dietary data much more manageable for researchers and much less burdensome for research participants, especially for those with cognitive challenges. DietID software relies on a clear and easy to use interface, which presents the user with a few foundational questions about their age, weight, height, and existing dietary restrictions (e.g., food allergies, religious restrictions). After collecting these baseline data, DietID utilizes a visual, picture-based system wherein users are asked to select images that most look like their average diet (dietary pattern). This visual approach is designed to evoke a sense of connection between the user and the foods they are seeing. The user is presented with two imagined diets, each made up of numerous images based on the foundational questions, in order to set a baseline. Then the participant is asked which set of images is most similar to their current diet, similar to an eye exam. Once an image is chosen, DietID recalibrates and produces two new sets of images with slightly varied diets (e.g., swapping butter for oil or whole wheat bread for white bread). This allows many diet permutations to be explored, with the goal of identifying a dietary pattern that is close to the user’s average intake as is possible. Once the final dietary pattern has been established, DietID utilizes a large database to generate a report of the approximated daily intake of vitamins, minerals, other nutrients, and key nutritional indices, i.e. the Healthy Eating Index (HEI). This report allows for direct comparison with dietary recommendations and provides the name of a dietary pattern to which the intake most closely aligns, e.g. the MedDiet. Participants in this study will have multiple ways to complete DietID: independently using their own devices (e.g., computers or tablets), with the assistance of a caregiver, or the assistance of members of the research team depending on their level of comfort with technology and cognitive impairment.

DietID was chosen as one of the key data-collecting methods for a myriad of reasons including its concise format, accurate results, and accessibility for the study population. The web-based, visual nature of DietID avoids the cognitive burdens of long-form surveys, dietary interviews, and creates a more approachable assessment that yields more accurate results (Rolstad et al., 2011). The accuracy of the software was also important to consider when implementing DietID, since image-based questions can be limiting and not reflective of all possible dietary patterns. Recent and thorough analyses of the software have shown that DietID’s non-invasive, yet rapid approach is a legitimate substitute for other diet-logging methods, such as the 24-hour dietary recall or FFQ (Radtke et al., 2023). Of note, the dietary data captured by DietID are intended to reflect short-term dietary intake, specifically dietary patterns over a 30-day reference period (Turner-McGrievy et al., 2021).

### Readiness for Change Survey

After a robust literature search for a survey that mapped stages of nutritional change, we found that there was no survey that fit our study objectives. However, there were multiple surveys which mapped stages of change for quitting smoking, alcohol use, physical activity and utilization of psychotherapy. We modeled our novel survey on the validated URICA, because of its reliable scoring mechanism that mapped survey responses to the appropriate stage of change.

To effectively measure the readiness to change and nutritional habits of our participants, we developed a 32-question survey based on the TTMC and URICA. We chose this model to capture the stage of change at which participants were regarding their desire to improve their nutritional habits. The wording of each statement on the URICA was adapted to discuss nutritional behaviors and the wording was conserved to resemble the original validated URICA survey as much as possible. For example, the first question was changed from, “as far as I’m concerned, I don’t have any problems that need changing” to “as far as I’m concerned, I don’t have any nutritional habits that need changing” (Heather et al., 1993; Henderson et al., 2004; McConnaughy et al., 1983). A five-point Likert scale was utilized for each survey statement. We named our survey, “Readiness to Change Nutritional Habits” (RCNH). Though our novel survey closely resembles the validated URICA, since it is adapted to fit our question, it is not validated and we hope to utilize the data we obtain from this protocol to aid in validation of this survey.

To refine the initial draft of the RCNH, we utilized both expert review and cognitive interviewing based on the six survey design principles described by Corr et al. (2023). Expert review consisted of sending the survey to two survey design experts who reviewed the draft survey for 1) adherence to the aims of the study and 2) question clarity. These expert reviewers were both well published in survey design and recognized as experts in the field; both of these experts are external to this particular study and have no other role in the design of instruments, data collection, of analysis. The final stage of the process was cognitive interviewing. This involved recruiting five individuals who resembled the study population. To achieve this purpose, three healthy control participants and two participants with mild cognitive impairment were recruited to discuss the draft instrument. Each participant was given the survey and asked to read each question out loud to the researchers. A think-aloud approach was utilized so that participants could state their thoughts on each survey item, ask questions, and provide recommended language and improvements (Corr et al., 2023). Each of these cognitive interviews generally lasted 30-60 minutes. After this process was completed, the suggested changes made by participants were added to the survey to enhance clarity and language use. This process was intended to make sure the survey wording was clear and understandable, therefore to aid in this process we opted for participants resembling the healthy control and mild cognitively impaired group. We omitted performing cognitive interviewing with dementia patients as the individualswe identified were not able to undertake the rask of survey editing due to the cognitive load and attention it required. The final draft of the survey is now part of the bMicrobiome Study. This survey data will be analyzed in conjunction with the gut microbiome profiling and dietary assessment data to explore the link between microbiome composition, dietary patterns, and cognitive health.

### Statistical Analyses

We will employ descriptive statistics to summarize demographic and clinical characteristics of participants across the three study groups. Correlational analyses (Pearson of Spearman’s Rho, depending on the distribution of participants) will be used to examine associations between readiness to change scores, dietary patterns, BOCA results, and microbiome diversity measures. Finally, we anticipate running exploratory regressions to determine if readiness to change may predict dietary patterns and microbiome diversity while adjusting for possible confounders (e.g., age, sex, cognitive status).

#### Hypothesis

This study hypothesizes that participants with greater readiness to change will demonstrate healthier eating patterns as reflected in higher Healthy Eating Index (HEI) scores from DietID. Additionally, we expect that greater gut microbiome diversity as measured by EzBiome reports will reflect a lower degree of cognitive decline.

## Discussion

In this sub-study, we specifically aimed to investigate how participants’ nutrition readiness to change influences their implementation of dietary behavior changes and the potential impact on gut microbiome composition and cognitive markers. By combining both objective data from microbiome analysis and subjective measures such as the RCNH survey, this approach allows for a deeper understanding of how individual readiness may shape dietary habits, ultimately affecting microbiome diversity and function, which are essential for cognitive health.

The findings of this sub-study could have significant implications for both scientific understanding and clinical practice in cognitive health. By exploring the relationship between gut microbiome composition and cognitive decline, and considering participants’ readiness to change dietary behaviors, this sub-study highlights the importance of intention to make behavior change as a factor that may affect health outcomes. If a strong association between nutrition readiness to change and microbiome composition is observed, it may suggest that personalized dietary interventions informed by both microbiome data and behavioral readiness could enhance adherence and efficacy in managing cognitive health.

With regard to clinical practice and public health interventions, an established link between microbiome composition and cognitive decline could lead to development of clinical guidelines that assess and manage cognitive health through gut microbiome profiling and gut microbiome directed interventions. Gut microbiome profiling could become part of routine evaluations for patients at risk of cognitive decline, allowing for early intervention and the possibility of prevention. Such assessments could be accompanied by personalized dietary recommendations, and lifestyle modifications designed to promote a healthy microbiome, potentially slowing the progression of cognitive decline. For example, such lifestyle modifications may include recommendations to increase fiber intake through fruits, vegetables, and whole grains; incorporating more plant-based foods; and reducing the consumption of highly processed foods. These approaches have all been recommended for supporting a diverse gut microbiome and may present useful recommendations for individuals based on their individual gut microbiome (Ayakdaş & Ağagündüz, 2023; Khavandegar et al., 2024; Nagpal et al., 2019).

Furthermore, assessing readiness to change offers valuable insights for designing patient-centered-interventions and may be useful in clinical practice to help generate a personalized nutritional and lifestyle change plan. Assessing intentions to make behavioral change is a difficult task; however, the TTMC provides the ability to effectively classify patients into a particular stage of change. With this classification, personalized dietary and lifestyle recommendations can be made that differ based on individual stage of change. For instance, individuals in precontemplation or contemplation stages may require educational support and motivational interviewing, whereas those in the action or maintenance stages might benefit more from specific dietary recommendations aligned with their microbiome profile. This tailored approach could improve adherence to lifestyle changes, potentially enhancing gut health and delaying cognitive decline in at-risk populations.

This study’s findings could pave the way for a personalized approach to cognitive health based on an individual’s unique microbiome profile, baseline cognitive health, readiness to make change, and current dietary pattern. The combination of these methodologies could yield a nuanced picture of how behavioral readiness impacts dietary implementation, which in turn may influence microbiome composition/function and related health outcomes.

### Strength and Limitations

This study design includes several strengths that help adequately answer the research question. First, the use of both objective and subjective data enhances the robustness of the findings by limiting potential self-reporting bias, which is a common limitation of survey-based research. Second, the partnership with EzBiome, the HIVE Lab, and NIST allows the use of next generation metagenomics to measure microbiome speciation and diversity and to infer function with independent verification. Third, the use of DietID for dietary pattern assessment provides an accurate dietary assessment that strongly correlates with traditional methods such as FFQs and even novel approaches such as measuring biomarkers, giving us more confidence in the objectivity of the dietary pattern data. Lastly, RCNH will be able to capture changes to participants’ readiness to eat healthier throughout the study period. This survey will be able to answer whether in-depth knowledge of one’s own microbiome leads to any change in desire to make nutritional changes.

Some possible drawbacks to our approach include the use of test kits requiring self-collection of stool samples. This may pose a challenge to many study participants —though, there is no feasible better alternative to this method. In regards to the EzBiome microbiome report, it is important to note that this is for research purposes only at present, and what constitutes a “healthy” microbiome is not well-defined in the current literature. It is also important to note that the gut microbiome is unique like a fingerprint, meaning there are many, many “healthy” gut microbiomes. The behavioral suggestions in the report are not necessarily proven to change or improve gut microbiome health; though, most are considered general good advice, e.g. eating more plants (McDonald et al., 2018). Because participation required the use of a digital device (for DietID and RCNH), our sample may be biased towards individuals with greater technological literacy and higher socioeconomic status. This limitation may affect the generalizability of findings. (To mitigate these issues, we have study staff ready to assist participants in accessing and utilizing these tools. Further, while we attempted to conserve the language of the URICA, we did make some changes in that we are applying this to a specific aspect of behavior change (nutrition) instead of behavior change broadly. Therefore, this is not a validated survey, and validation would strengthen future survey research and is a logical next step. In addition, the current iteration of this survey aims to capture readiness to change at a macro level and does not discriminate what types of food the participant may be adopting. Therefore on its own this survey may be very broad and not able detect granular change, however this issue is mitigated in this particular study as the participants presumed nutrition goal is outlined for them in the personalized microbiome report. For example, if a p participant’s report shows they need eat more beans, we can infer they fill out the survey with regard to making that specific lifestyle change. Finally, we would be remiss if we did not acknowledge the critical role of informal caregivers in the support of individuals with cognitive impairment. In the context of this study, however, a person’s dietary habits may be shaped by household roles and gender norms in terms of food purchasing, preparation, and patient care (Atta-Konadu, 2011; Russell et al., 2007).

After completion of this study, we expect the results to demonstrate a correlation between the gut microbiome (composition / predicted function), dietary patterns, and readiness to change nutritional habits. We expect participants with a diet rich in fiber, polyunsaturated fats, and polyphenols, all of which are found in the MedDiet, to have greater microbiome diversity than participants with diets rich in red-meat and simple carbohydrates (Merra et al., 2020). We also expect to see participants who are further along in the stages of change model, as measured by our RCNH, to have healthier dietary patterns and, thus, less dysbiotic microbiomes. However, it is likely that cognitive deficit will play a role in RCNH and DietID results. It may also play a role with their understanding of the EzBiome report and its behavioral suggestions, which would be reflected by a lack of positive nutritional change throughout the study intervals. This may be mitigated somewhat by the inclusion of the caregiver.

### Next Steps

Building on the findings of this sub-study, future research should focus on validating the relationships between nutrition readiness to change, dietary behaviors, and microbiome composition in larger and more diverse populations. One priority is to refine and validate the RCNH survey specifically for dietary behavior change in populations with cognitive impairment. This could enable broader clinical application and allow for a more standardized approach to assessing readiness for dietary change in such populations.

Additionally, future studies should explore the potential long-term impacts of personalized dietary interventions based on microbiome composition and readiness to change. Implementing follow-up periods extending beyond six months could provide insights into the sustainability of dietary behavior changes and their ongoing influence on the microbiome-gut-brain axis. Moreover, examining how individual differences in cognitive status might affect adherence to dietary recommendations will help tailor interventions to meet the unique needs of patients across different stages of cognitive impairment.

Though this study does not assess Apolipoprotein E4 (APOE4) status, addition of APOE4 and other relevant biomarkers to a future iteration of this protocol could provide further insight into the relationship between the microbiome and cognitive decline. Studies show that APOE4 carriers have distinct microbiome phenotypes notably with depletion of protective bacteria (Faecalibacterium, Ruminococcus, and Butyricoccus) and increase in pro inflammatory bacteria (Alistipes and Bacteroides) which are associated with increase amyloid deposition (Wadop et al., 2025, Tran et al., 2019; Hou et al., 2021). Assessment of microbiome change over time compared between APOE4 carriers and non-carriers would be a worthwhile future step to further establish the significance of APOE4 in its relationship between dementia and the gut microbiome.

Integrating multi-omics approaches, such as metabolomics and transcriptomics, would offer deeper insights into the functional implications of microbiome shifts and how these may impact cognitive health. Finally, expanding the study to assess other behavioral factors, such as physical activity and sleep, would provide a more comprehensive picture of lifestyle modifications and their cumulative effect on both microbiome composition and cognitive outcomes. By addressing these areas, future research could strengthen the evidence base for personalized, microbiome-informed dietary strategies in promoting cognitive resilience.

## Conclusion

This sub-study demonstrates the potential for combining microbiome analysis with readiness to change assessments to inform personalized dietary interventions that may benefit cognitive health. By evaluating how participants’ readiness to adopt dietary changes influences gut microbiome composition and associated cognitive health markers, this research underscores the role of readiness to make behavior change in achieving health outcomes. The findings suggest that personalized interventions tailored to an individual’s microbiome profile and readiness for change could enhance adherence and potentially mitigate cognitive decline.

Our approach, which integrates microbiome sequencing, dietary assessment, and behavioral readiness, offers a promising framework for future studies aiming to promote cognitive resilience through lifestyle interventions. Ultimately, this study lays the groundwork for more targeted, patient-centered strategies in cognitive health care, supporting the growing body of evidence on the gut-brain axis and its implications for neurodegenerative conditions.

## Data Availability

All data produced in the present study are available upon reasonable request to the authors.

## Author Contributions

Conceptualization: K.S., P.G.C., and L.A.F.; Data Curation: K.S., A.S., and A.A.; Methodology: P.G.C. and L.A.F.; Formal Analysis: K.S., A.S., A.A. and P.G.C.; Visualizations: K.S., A.S., A.A., and L.A.F.; Writing – Original Draft: K.S., A.S. A.A., P.G.C., and L.A.F.; Writing – Review and Editing: K.S., P.G.C., and L.A.F.; Supervision: P.G.C. and L.A.F.; Funding Acquisition: L.A.F.

## Acknowledgments

We would like to gratefully acknowledge Alison Warren, Nur Hasan, Abdelmohsen Al Qalam, Mina Farah, Michaela Karam, Katherine Rangoussis, Mikhail Kogan, members of the team leading the launch of the main study, without which this sub-study would not be possible.

## Ethical Approval

This study was reviewed and approved by the George Washington University Institutional Review Board (Approval #: NCR234792) and all participants completed a written informed consent.

## Informed Consent

All participants completed a signed informed consent prior to enrolling in the study.

## References

Atta-Konadu, E., Keller, H. H., & Daly, K. (2011). The food-related role shift experiences of spousal male care partners and their wives with dementia. Journal of Aging Studies, 25(3), 305–315

Ayakdaş, G., & Ağagündüz, D. (2023). Microbiota-accessible carbohydrates (MACs) as novel gut microbiome modulators in noncommunicable diseases. Heliyon, 9(9), e19888. 10.1016/j.heliyon.2023.e19888

Barber, C., Mego, M., Sabater, C., et al. (2021). Differential effects of Western and Mediterranean-type diets on gut microbiota: A metagenomics and metabolomics approach. Nutrients, 13, 2638. 10.3390/nu13082638

Bernstein, A. M., Rhee, L. Q., Njike, V. Y., & Katz, D. L. (2023). Dietary assessment by pattern recognition: A comparative analysis. Current Developments in Nutrition, 7(10), 101999. 10.1016/j.cdnut.2023.101999

Corr, P. G., Aly, R., & Artino, A. (2023). Survey research for health professionals. In E. Rees, A. Ledger, & K. Walker (Eds.), Starting research in clinical education (pp. 183–191). Wiley Education Services.

Dansinger, M. L., Breton, G. L., Joly, J. E., Rhee, L. Q., & Katz, D. L. (2023). Rapid, digital dietary assessment in association with cardiometabolic biomarkers. American Journal of Health Promotion. 10.1177/08901171231156513

Ferreiro, A. L., Choi, J., Ryou, J., et al. (2023). Gut microbiome composition may be an indicator of preclinical Alzheimer’s disease. Science Translational Medicine, 15(700), eabo2984. 10.1126/scitranslmed.abo2984

Godny, L., Reshef, L., Sharar Fischler, T., et al. (2022). Increasing adherence to the Mediterranean diet and lifestyle is associated with reduced fecal calprotectin and intra-individual changes in microbial composition of healthy subjects. Gut Microbes, 14, e2120749. 10.1080/19490976.2022.2120749

Guinane, C. M., & Cotter, P. D. (2013). Role of the gut microbiota in health and chronic gastrointestinal disease: Understanding a hidden metabolic organ. Therapeutic Advances in Gastroenterology, 6(4), 295–308. 10.1177/1756283X13482996

Harding, S. L., & Bishop, J. (2022). The gut microbiome, mental health, and cognitive and neurodevelopmental disorders: A scoping review. The Journal for Nurse Practitioners, 18(7), 719–725. 10.1016/j.nurpra.2022.04.019

Hashimoto, K. (2023). Emerging role of the host microbiome in neuropsychiatric disorders: Overview and future directions. Molecular Psychiatry, 28, 3625–3637. 10.1038/s41380-023-02287-6

Heather, N., Rollnick, S., & Bell, A. (1993). Predictive validity of the Readiness to Change Questionnaire. Addiction, 88(12), 1667–1677. 10.1111/j.1360-0443.1993.tb02042.x

Henderson, M. J., Saules, K. K., & Galen, L. W. (2004). The predictive validity of the University of Rhode Island Change Assessment Questionnaire in a heroin-addicted polysubstance abuse sample. Psychology of Addictive Behaviors, 18(2), 106–112. 10.1037/0893-164X.18.2.106

Hirschberg, S., Gisevius, B., Duscha, A., & Haghikia, A. (2019). Implications of diet and the gut microbiome in neuroinflammatory and neurodegenerative diseases. International Journal of Molecular Sciences, 20(12), 3109. 10.3390/ijms20123109

Hou, M., Xu, G., Ran, M., Luo, W., & Wang, H. (2021). APOE-ε4 carrier status and gut microbiota dysbiosis in patients with Alzheimer disease. Frontiers in Neuroscience, 15, 619051. 10.3389/fnins.2021.619051

Katz, D. L., Rhee, L. Q., Katz, C. S., Aronson, D. L., Frank, G. C., Gardner, C. D., Willett, W. C., & Dansinger, M. L. (2020). Dietary assessment can be based on pattern recognition rather than recall. Medical Hypotheses, 140, 109644. 10.1016/j.mehy.2020.109644

Khavandegar, A., Heidarzadeh, A., Angoorani, P., et al. (2024). Adherence to the Mediterranean diet can beneficially affect the gut microbiota composition: A systematic review. BMC Medical Genomics, 17, 91. 10.1186/s12920-024-01861-3

Liang, X., Fu, Y., Wen-ting, C., et al. (2022). Gut microbiome, cognitive function and brain structure: A multi-omics integration analysis. Translational Neurodegeneration, 11, 1–14. 10.1186/s40035-022-00323-z

Manderino, L., Carroll, I., Azcarate-Peril, M., et al. (2017). Preliminary evidence for an association between the composition of the gut microbiome and cognitive function in neurologically healthy older adults. Journal of the International Neuropsychological Society, 23(8), 700–705. 10.1017/S1355617717000492

McConnaughy, E. N., Prochaska, J. O., & Velicer, W. F. (1983). Stages of change in psychotherapy: Measurement and sample profiles. Psychotherapy, 20, 368–375

McDonald, D., Hyde, E., Debelius, J. W., et al. (2018). American Gut: An open platform for citizen science microbiome research. mSystems, 3(3), e00031–18. 10.1128/mSystems.00031-18

Merra, G., Noce, A., Marrone, G., et al. (2020). Influence of Mediterranean diet on human gut microbiota. Nutrients, 13(1), 7. 10.3390/nu13010007

Nagpal, R., Shively, C. A., Register, T. C., Craft, S., & Yadav, H. (2019). Gut microbiome-Mediterranean diet interactions in improving host health. F1000Research, 8, 699. 10.12688/f1000research.18992.1

Prochaska, J. O., & DiClemente, C. C. (1992). Stages of change in the modification of problem behaviors. Progress in Behavior Modification, 28, 183–218.

Prochaska, J. O., & Velicer, W. F. (1997). The transtheoretical model of health behavior change. American Journal of Health Promotion, 12(1), 38–48. 10.4278/0890-1171-12.1.38

Quigley, E.M.M/ (2017). Microbiota-Brain-Gut Axis and Neurodegenerative Diseases. Current Neurology and Neuroscience Reports;17(12). doi:10.1007/s11910-017-0802-6

Radtke, M. D., Chodur, G. M., Bissell, M. C. S., Kemp, L. C., Medici, V., Steinberg, F. M., & Scherr, R. E. (2023). Validation of Diet ID™ in predicting nutrient intake compared to dietary recalls, skin carotenoid scores, and plasma carotenoids in university students. Nutrients, 15, 409. 10.3390/nu15020409

Raihan, N., & Cogburn, M. (2024). Stages of change theory. In StatPearls. StatPearls Publishing. https://www.ncbi.nlm.nih.gov/books/NBK556005/

Rejeski, J. J., Wilson, F. M., Nagpal, R., et al. (2022). The impact of a Mediterranean diet on the gut microbiome in healthy human subjects: A pilot study. Digestion, 103, 133–140. 10.1159/000519445

Rolstad, S., Adler, J., & Rydén, A. (2011). Response burden and questionnaire length: Is shorter better? A review and meta-analysis. Value in Health, 14(8), 1101–1108. 10.1016/j.jval.2011.06.003

Russell, R. (2007). Men doing “women’s work:” Elderly men caregivers and the gendered construction of care work. Journal of Men’s Studies, 15(1), 1–18. 10.3149/jms.1501.1

Shaffer, J. A. (2013). Stages-of-change model. In M. D. Gellman & J. R. Turner (Eds.), Encyclopedia of behavioral medicine. Springer. 10.1007/978-1-4419-1005-9_1180

Silva, Y. P., Bernardi, A., & Frozza, R. L. (2020). The role of short-chain fatty acids from gut microbiota in gut-brain communication. Frontiers in Endocrinology, 11, 25. 10.3389/fendo.2020.00025

Sinha, A. K., Laursen, M. F., Brinck, J. E., et al. (2024). Dietary fibre directs microbial tryptophan metabolism via metabolic interactions in the gut microbiota. Nature Microbiology. 10.1038/s41564-024-01737-3

Sutton, J. (2022). How to assess and change readiness for change. PositivePsychology.com. https://positivepsychology.com/readiness-for-change/

Ticinesi, A., Nouvenne, A., Cerundolo, N., Parise, A., Mena, P., & Meschi, T. (2024). The interaction between Mediterranean diet and intestinal microbiome: Relevance for preventive strategies against frailty in older individuals. Aging Clinical and Experimental Research, 36(1), 58. 10.1007/s40520-024-02707-9

Tran, T. T. T., Corsini, S., Kellingray, L., Hegarty, C., Le Gall, G., Narbad, A., Müller, M., Tejera, N., O’Toole, P. W., Minihane, A.-M., & Vauzour, D. (2019). APOE genotype influences the gut microbiome structure and function in humans and mice: Relevance for Alzheimer’s disease pathophysiology. FASEB Journal, 33(7), 8221–8231. 10.1096/fj.201900071R

Turner-McGrievy, G., Hutto, B., Bernhart, J. A., & Wilson, M. J. (2021). Comparison of the Diet ID platform to the automated self-administered 24-hour (ASA24) dietary assessment tool for assessment of dietary intake. Journal of the American College of Nutrition, 40(3), 242–264.

Vyshedskiy, A., Netson, R., Fridberg, E., Jagadeesan, P., Arnold, M., Barnett, S., Gondalia, A., Maslova, V., de Torres, L., Ostrovsky, S., Durakovic, D., Savchenko, A., McNett, S., Kogan, M., Piryatinsky, I., & Gold, D. (2022). Boston cognitive assessment (BOCA) - a comprehensive self-administered smartphone- and computer-based at-home test for longitudinal tracking of cognitive performance. BMC neurology, 22(1), 92. 10.1186/s12883-022-02620-6

Wadop, Y. N., Bernal, R., Njamnshi, W. Y., et al. (2025). Altered gut microbiota mediates the association between APOE genotype and amyloid-β accumulation in middle-aged adults. Journal of Neurology, 272(10), 670. 10.1007/s00415-025-13412-6

Warren, A., Nyavor, Y., Zarabian, N., Mahoney, A., & Frame, L. A. (2024). The microbiota-gut-brain-immune interface in the pathogenesis of neuroinflammatory diseases: A narrative review of the emerging literature. Frontiers in Immunology, 15, 1365673. 10.3389/fimmu.2024.1365673

Wolf, P. G., Byrd, D. A., Cares, K., Dai, H., Odoms-Young, A., Gaskins, H. R., Ridlon, J. M., & Tussing-Humphreys, L. (2022). Bile acids, gut microbes, and the neighborhood food environment—A potential driver of colorectal cancer health disparities. mSystems, 7, e01174–21. 10.1128/msystems.01174-21

Zhang, P. (2022). Influence of foods and nutrition on the gut microbiome and implications for intestinal health. International Journal of Molecular Sciences, 23(17), 9588. 10.3390/ijms23179588

